# Who Engages in a National Sports Club-Based Community Walking Initiative? Reach and Participant Characteristics from the Ireland Lights Up Programme

**DOI:** 10.64898/2026.06.18.26355653

**Authors:** Nicola Briggs, Barry Lambe, Niamh Murphy, Noel Richardson, Aisling McGrath

**Affiliations:** Department of Sport and Exercise Science, South East Technological University, Waterford, Ireland; National Centre for Men’s Health, South East Technological University, Carlow, Ireland

**Keywords:** physical activity, walking, reach, community, health promoting sports clubs, inclusion

## Abstract

Sports clubs are gaining recognition as valuable settings for population health promotion, extending impact beyond traditional sporting populations. Ireland Lights Up (ILU) is a nationwide community walking initiative in which Gaelic Athletic Association (GAA) clubs provide floodlit walking opportunities during winter to engage communities in physical activity (PA). However, there is limited evidence on who such initiatives reach and whether they enage diverse population groups. To address this gap, a mixed methods approach was utilised. Quantitative data was obtained through cross-sectional questionnaires distributed to participants (n=1226) through club networks over a six-week period. Qualitative data was collected through snapshot interviews (n=30) with participants during site visits and triangulated to understand drivers of participation. Findings indicated that most participants were female (76.5%), white (99.5%) and lived in rural areas (80%). Participants had a mean age of 47 years (SD ±11.01 with the majority (84%) as sports club members. Participants reported high levels of health and wellbeing, with a substantial proportion meeting PA guidelines (48.1%). Single and younger participants were more likely to feel lonely compared to married and older participants (P≤0.05). Qualitative findings highlighted three main participation drivers: safety, social interaction and club walking tracks. ILU engages large numbers of participants in PA nationally and successfully engages its target population, including substantial numbers from traditionally underserved rural communities, by employing an innovative approach that addresses seasonal barriers to PA participation. To further maximise reach, community-based PA programmes require research that includes attention to implementation factors such as participant acceptability.

## BACKGROUND

Since designating physical inactivity as a global pandemic, the call for effective and scalable strategies to increase population physical activity (PA) has escalated [1, 2]. While traditional efforts to increase health enhancing PA focused on individual behaviour change, there is now greater emphasis on the broader social, environmental, and policy contexts that shape population health [3]. As a result, the settings-based approach has emerged as a key strategy to support sustainable health outcomes [4]. As a settings-based health promotion (HP) measure, walking groups have proven to be an effective way to enhance health, drawing large numbers of participants and maintaining low attrition rates [5, 6]. Walking is a low-cost activity that carries minimal risk of injury and requires little technical ability [7]. Evidence from systematic reviews indicates that community-based group walking programmes are effective in increasing PA levels and promoting overall health [5, 8, 9]. Examples include 10,000 Steps Rockhampton [10] and Active Launceston in Australia [11] which evidenced significant project reach and may act as models for implementing community-wide interventions. Walking groups show particular promise in engaging certain demographic groups that face barriers to PA, such as women, older adults and those living in rural areas [12–14].

Increasingly, sports clubs are seen as effective settings for health promotion, as they can extend their influence beyond traditional approaches and promote health-enhancing activities [15, 16]. Sports clubs are positioned to engage a broader audience beyond more competitive or elite athletes, including groups from diverse socioeconomic backgrounds and life stages [17]. Alongside their primary focus on elite performance, community sports clubs also operate as social entities, playing a vital role in promoting social health and supporting the physical and psychosocial wellbeing of the communities they serve [18, 19]. Thus, by facilitating outdoor spaces for walking interventions, sports clubs can be an important setting for HP [7, 20]. However, evidence of large-scale, sustainable community walking initiatives is limited, particularly in the sports club setting [2, 14].

An example of health promoting sports clubs (HPSC) is the Gaelic Athletic Association (GAA), Ireland’s largest amateur sporting organisation, which has a deep-rooted community presence alongside the promotion of traditional Irish sport [21]. According to official data [22], there are 1610 registered GAA clubs in Ireland, operating from 1,597 unique club facilities. The current GAA membership stands at an estimated 550,186 members, representing 7.85% of the entire Irish population [22]. However, this figure likely underestimates the true reach of the organisation, as many additional community members engage with local clubs in a voluntary or informal capacity without holding official membership. The GAA is strongly tied to Irish identity and community life, particularly in rural areas, where clubs are organised around parish and county structures that foster strong local affiliation and rivalry. While this cultural embeddedness provides a powerful platform for community mobilisation and engagement, it may also present challenges in extending reach beyond existing networks, particularly among individuals who do not identify with the organisation or its cultural associations.

A GAA hosted walking initiative, ‘Ireland Lights Up’ (ILU), was introduced in 2018 through partnership with Get Ireland Walking (a national initiative of Sport Ireland), Healthy Ireland (a government-led framework for action to improve health and wellbeing in Ireland) and Operation Transformation (a cross-media health programme on RTÉ, Ireland’s national broadcaster). Healthy Ireland allocates government funding to allow eligible sports clubs to upgrade their walking tracks to encourage more people to be physically active. The ILU initiative is designed to reach the general public including club members and non-members, communicated through GAA Head Office, local clubs and national media outlets. Using a simple premise, ILU encourages GAA clubs across Ireland to open their grounds to local communities and turn on pitch side floodlights for volunteer-led weekly walks over a six-week period during the months of January and February when daylight is limited. The current initiative did not start from a green field site but sought to capitalise on an existing and well-established health promotion and partnership model, the Healthy Club Programme (HCP) where the GAA seeks to support communities to achieve health and wellbeing goals [23, 24]. As part of the HCP, the ‘Irish Life (a financial services group) Every Step Counts Challenge’ has run concurrently with ILU since 2021, with clubs challenged to reach a collective step target measured via the MyLife app. In 2024, it was estimated by the GAA that a national average of 40,000 people took part in ILU each week [25]. Now operating at scale across all 32 counties of Ireland annually, the initiative has shown preliminary benefits including increased PA, improved general health and wellbeing, and enhanced social connectedness among participants [26]. However, as a fast-growing initiative emerging from the real-world, there is a lack of evidence regarding its reach and adoption across both the individual and club level.

ILU is a flagship PA initiative that clubs can choose for inclusion in the HCP, yet there is no data on who participates or why. This is important not only for ILU but also for designing future community or sports club-based walking interventions that wish to engage a representative cohort of the community through outreach with those who may not be typically affiliated with sport clubs like the GAA. Community-based initiatives are increasingly focused on engaging greater sections of the general population, particularly ‘not yet reached’ groups from lower socioeconomic backgrounds, ethnic minorities and rural populations [2, 27, 28]. Investigating who participates addresses a knowledge gap, particularly when aiming to engage underreached populations. To fully understand why public health interventions work, researchers must examine the characteristics of people for whom an intervention was effective, as well as those for whom it was less so. These differences may stem from the way intervention strategies were delivered, or from the context in which they were implemented [29]. However, evaluations often omit these contextual factors, which are particularly important when attempting to reach underserved populations [30]. This can only be determined by examining interventions through both a qualitative and a quantitative lens. Equity should not be just about ‘opening the gates’ of club grounds without intentional strategies as participation will still reflect existing membership profiles [31, 32].

In evaluating such initiatives, the Reach, Effectiveness, Adoption, Implementation and Maintenance (RE-AIM) framework [33] has been used as a tool for assessing multi-level factors involved in PA interventions [2, 34]. It has been applied to a range of programmes across various settings such as workplaces [35], schools [36], disadvantaged areas [37] and rural communities [20]. The application of the framework in this study has included a focus on the concept of ‘reach’, examining the factors that encourage individuals to participate and the elements that influence overall participation. The purpose of this study is to assess the participant reach and club adoption of ILU by examining the demographic characteristics that influence participation, identifying who is not reached, and exploring how the cultural context of the GAA, the physical environment and the nature of walking as the form of PA shapes patterns of participation.

## METHODS

This research was part of a broader implementation study evaluating the ILU initiative. The wider study utilises a mixed methods implementation science design, fully described in prior work [38], which integrates the systematic planning frameworks: RE-AIM [33] and the Practical, Robust Implementation and Sustainability Model (PRISM) [34]. This design allows for an iterative evaluation of reach, effectiveness, adoption, implementation and maintenance. In this context, reach is best described as participation rates within a target population and the demographic characteristics of participants versus non-participants [39]. Adoption applies at the organisational level and refers in this study to the percentage of GAA clubs that participate in this particular initiative [40].

This study primarily relates to the reach and adoption components within the RE-AIM framework. The programme is continuing and effectiveness, implementation and maintenance will be discussed in future papers. Utilising a mixed methods design, data was collected using cross-sectional surveys (n=1226) and semi-structured snapshot interviews (n=30) conducted during site visits. “Snapshot interviews” refer to brief, semi-structured interviews conducted in situ, that is, at the time and place of participant engagement with the initiative, to capture immediate perceptions, experiences and contextual insights in real time that can be lost in retrospective interviews [41]. The study received ethical approval from the Health Sciences Research Ethics Committee (SETU/HSREC/23/24/010) and is registered with the ‘International Standard Randomised Controlled Trial Number’ registry (ISRCTN 14693503).

### Data Collection

For this study, non-random convenience sampling was used to recruit a diverse sample of clubs, participants and organisers across the GAA. A list of clubs was obtained from the GAA and each club was contacted to gauge their interest in participating in the observational component of the study. Data were collected during two periods of the initiative in 2024 and 2025. Quantitative data consisted of cross-sectional surveys from ILU participants (n=1226) along with ILU registration records from the GAA. The participant demographic characteristics survey was administered online through the electronic data collection platform, Qualtrics and disseminated through GAA gatekeepers to all participating clubs (n=784). This one-time, cross-sectional survey aimed to characterise participants of ILU and contained 4 sections. The survey collected demographic information (e.g., age, gender, occupation), GAA club membership details (e.g., affiliation, belonging), and ILU participation data (e.g., frequency). To examine patterns of ongoing engagement, a question assessed participation in ILU during previous years. Participant PA levels were measured using the Single Item Measure [42] and followed the standard PA guidelines in Ireland which recommends at least 30 minutes of moderate-intensity activity a day, five days a week [43]. Validated measures assessed strength training levels [44], sense of belonging [45], loneliness using the UCLA 3-item Loneliness Scale [46], life satisfaction [47], and self-reported health status [48]. Participants were also asked for feedback on ILU satisfaction levels by rating on a scale of 0-10 and to explain the reason for the rating.

Qualitative data consisted of semi-structured snapshot interviews (n=30) conducted with participants during site visits. To ensure diversity, clubs were selected through consultation with the GAA Community and Health team. Each selected club was visited once by a researcher where snapshot interviews, and systematic field notetaking were conducted. Consent was obtained in advance from club organisers. Field notes were completed using a printed observational tool along with a small notebook and a Dictaphone. The researcher adopted a two-phase observational approach, beginning with observation from a distance followed by observation through participation to capture both environmental dynamics and in-situ social interactions. Semi-structured snapshot interviews were conducted and voice recorded using a Dictaphone during site visits. During the walks, the primary researcher approached groups of two or three participants at random, providing an explanation of the research purpose. The interviews commenced after informed consent was obtained. The interview schedule was informed by the RE-AIM framework and included questions about level of engagement by the participant, how ILU was promoted by the club and reasons for participation.

### Data Analysis

As this was a mixed methods study, reporting was guided by the Good Reporting of a Mixed Methods Study (GRAMMS) framework [49], which provides criteria for the transparent description of design, integration, interpretation, and limitations. Statistical Packages for the Social Sciences (SPSS V25) was utilised to analyse questionnaire data. Descriptive statistics were calculated for all baseline characteristics. Group differences at baseline were assessed using independent-samples t tests and post-hoc analysis depending on the type of variable. Where appropriate, bivariate analyses were conducted to examine the strength and direction of associations between variables. Data relating to participation (attendance records, participant and club numbers who participated versus numbers eligible) were cross-referenced to assess reach.

The RE-AIM and PRISM framework-driven approach was used to analyse qualitative data. Interviews were transcribed verbatim and in accordance with recommendations by the National Cancer Institute [50] on qualitative research in implementation science, a hybrid approach of thematic deductive and inductive analysis was applied. Thematic analysis was conducted by the primary researcher and subsequently verified through discussion with two additional team members, with findings further strengthened by triangulating quantitative and qualitative data across a series of full research team discussions. The research team comprised of academics with backgrounds in health promotion, exercise science, and community health. As researchers embedded within this field, the team held an inherent interest in the success of community-based PA interventions, which may have shaped their interpretive lens. To mitigate this, ongoing critical reflection was maintained throughout analysis, including regular team discussions, reflexive journaling during fieldwork, and close attention to participant accounts that complicated or contradicted initial assumptions.

Data collection was conducted by the primary researcher who was not directly involved in programme delivery but did participate in the walks during site visits. As such, the team acknowledges that the researcher identity, including professional roles, gender, and institutional affiliations may have influenced how participants presented their experiences. An interpretivist standpoint was adopted, treating participant accounts as situated and subjective rather than fixed representations of a single truth. Analysis prioritised participant meaning-making, with iterative returns to the data to ground interpretations in participants’ own words and experiences.

## RESULTS

### Quantitative Results

#### Ireland Lights Up Adopting Club Characteristics

Analysis of 2024 ILU registration records identified 784 distinct clubs as participating, equating to 48.7% of the 1,610 clubs registered in Ireland and representing close to half of all eligible clubs nationwide. The majority of the clubs’ facilities (n=619, 79%) included dedicated walking tracks, while the remaining clubs (n=165, 21%) utilised public lit routes in the vicinity of their grounds. The regional breakdown of registered clubs by province was found to be highest in Leinster (n=358, 34.45%), followed by Munster (n=282, 27.14%), Ulster (n=269, 25.89%) and Connacht (n=128, 12.31%). According to the GAA, in 2023, 56% of clubs (n=211) involved in the Irish Life Healthy Club Programme also participated in ILU.

In 2024, ILU engaged an estimated 40,000 participants per week across its six-week duration [25], indicating substantial national scale. However, as ILU is open to both club members and the wider community, a precise overall reach percentage cannot be calculated because the total eligible population is unknown. The majority of respondents who participated in ILU were affiliated with a local club (n = 1009, 88.2%), suggesting strong internal reach within existing club networks. Participants also appeared to be highly local, with 83.6% living within 5 km of a club and only 0.4% living more than 20 km from a GAA facility. Overall, ILU participants had taken part for an average of 1.56 years (SD ± 0.497), indicating repeat engagement across programme cycles.

#### Ireland Lights Up Participant Characteristics

A total of 1266 respondents took part in the demographic survey. As shown in **Table 1** below, the majority of participants were female (n=823, 76.5%). The sample was predominantly white (n=1069, 99.5%), with a small minority identifying as black (n=2, 0.2%), Asian (n=1, 0.1%), or from other/mixed backgrounds (n=2, 0.2%). The mean age of participants was 46.9 years (n=1061, SD±11.01), with a similar distribution of males (47.37, SD±10.867) and females (46.94). The majority of participants were aged 35–54 (n=695, 65.7%), with most participants having a third level or postgraduate education (n=859, 80.1%). A high proportion were employed either full or part time (n=861, 80.2%). Most participants were married, cohabiting or in a relationship (n=918, 85.6%). Participants reported living in predominantly rural locations (n=913, 80.1%), with only 19.9% (n=227) in urban locations.

**Table 1.** ILU participant demographic characteristics.

| Variable (measurement used) | All<br>N (%) | Male<br>N (%) | Female<br>N (%) |
| --- | --- | --- | --- |
| **Gender (multiple choice)** |  | 253 (23.5) | 823 (76.5) |
| **Ethnicity (multiple choice)** |  |  |  |
| White (Irish, any other White background) | 1069 (99.5) | 250 (99.2) | 819 (99.6) |
| Black (Irish, any other Black background) | 2 (0.2) | 1 (0.4) | 1 (0.1) |
| Asian (Irish, any other Asian background) | 1 (0.1) | 0 | 1 (0.1) |
| Other backgrounds, including mixed | 2 (0.2) | 1 (0.4) | 1 (0.2) |
| **Education Level (multiple choice)** |  |  |  |
| Primary education only | 8 (0.7) | 4 (1.6) | 4 (0.5) |
| Secondary education | 206 (19.2) | 73 (29) | 133 (16.2) |
| Third level education | 548 (51.1) | 123 (48.8) | 425 (51.8) |
| Postgraduate education | 311 (29) | 52 (20.6) | 259 (31.5) |
| **Employment Status (multiple choice)** |  |  |  |
| Employed full or part time | 861 (80.2) | 222 (88.1) | 639 (77.6) |
| Not currently employed | 12 (1.1) | 2 (0.8) | 10 (1.2) |
| Looking after home/family | 85 (7.9) | 3 (1.2) | 82 (10.0) |
| Retired | 85 (7.9) | 14 (5.6) | 71 (8.6) |
| Student | 21 (2) | 7 (2.8) | 14 (1.7) |
| Unable to work due to disability | 6 (0.6) | 3 (1.2) | 3 (0.4) |
| Other | 5 (0.5) | 1 (0.4) | 4 (0.5) |
| **Relationship Status (multiple choice)** |  |  |  |
| Married/cohabiting | 870 (81.1) | 210 (83.0) | 660 (80.5) |
| Single | 114 (10.6) | 24 (9.5) | 90 (11.0) |
| In a relationship | 48 (4.5) | 11 (4.3) | 37 (4.5) |
| Separated/Divorced | 28 (2.6) | 8 (3.2) | 20 (2.4) |
| Other | 13 (1.2) | 0 | 13 (1.6) |
| **Geographical Location (multiple choice)** |  |  |  |
| Urban | 214 (19.9) | 53 (20.9) | 161 (19.6) |
| Rural | 859 (80.1) | 200 (79.1) | 659 (80.4) |
| **Current Sports Club Membership (multiple choice)** |  |  |  |
| Yes | 913 (84.9) | 241 (95.3) | 672 (81.8) |
| No | 122 (11.3) | 8 (3.2) | 114 (13.9) |
| Past member | 40 (3.7) | 4 (1.6) | 36 (4.4) |
|  | **All (range)**<br>**±SD** | **Male** | **Female** |
| **Age in years (mean)** | 46.9 (18-80)<br>±11.01 | 47.37 | 46.94 |

#### Health and Wellbeing Characteristics

Nearly half of respondents (n=509, 48.1%) met the recommended PA guidelines of ≥150 minutes of moderate-intensity activity per week (see **Table** 2 below), compared with 41% in the general population [51]. Likewise, over half of participants (n=467, 53.6%) met the strength training guidelines of at least two days per week (M=2.04, SD±1.898). Over 89% (n=965) of respondents reported positive overall health, compared to the general population in Ireland at 75.8% [52].

**Table 2.** ILU participant health and wellbeing characteristics.

| Variable (measurement used) | All<br>N (%) | Male<br>N (%) | Female<br>N (%) |
| --- | --- | --- | --- |
| **Self-rated health (single question Likert scale)** |  |  |  |
| Very good to excellent | 965 (89.8) | 222 (87.8) | 743 (90.3) |
| Good | 101 (9.4) | 29 (11.4) | 72 (8.8) |
| Average to poor | 9 (0.8) | 2 (0.8) | 7 (0.9) |
| **Belonging (Likert scale assessing feelings of belonging)** |  |  |  |
| Agree to Strongly Agree | 1040 (96.8) | 237 (94) | 803 (97.7) |
| Disagree to Strongly Disagree | 34 (3.2) | 15 (6) | 19 (2.3) |
| **Close support (Likert scale assessing feelings of close support)** |  |  |  |
| Agree to Strongly Agree | 1017 (94.7) | 234 (92.9) | 783 (95.3) |
| Disagree to Strongly Disagree | 57 (5.3) | 18 (7.1) | 39 (4.7) |
| **Loneliness Score (UCLA three-item scale)** |  |  |  |
| Not Lonely | 793 (74.3) | 188 (75.2) | 605 (74.1) |
| Lonely | 274 (25.7) | 62 (24.8) | 212 (25.9) |
| **Meeting PA guidelines (calculated from the eight-point single item PA measure)** |  |  |  |
| Yes | 509 (48.1) | 124 (49.8) | 385 (47.6) |
| No | 549 (51.9) | 125 (50.2) | 424 (52.4) |
| **Meeting strength guidelines (calculated from the eight-point single item strength measure)** |  |  |  |
| Yes | 467 (53.6) | 132 (60.8) | 335 (51.2) |
| No | 404 (46.4) | 85 (39.2) | 319 (48.8) |
|  | **All (n)** | **Mean (range) ±SD** |  |
| **Life satisfaction (mean values on an 11-point scale)** | 1065 | 7.74 (1-10) ±1.448 |  |

In this sample, 74.3% (n=793) of participants were classified as ‘not lonely’, compared with 67.3% in the general population [52]. However, despite this positive indicator, as per UCLA, one quarter (n=274, 25.7%) of participants reported experiencing loneliness. Post hoc analysis confirmed a statistically significant difference in loneliness levels across age groups (f = 2.677, p = .021), with the 18-24 age group reporting significantly higher loneliness than the 65+ group (p = .010). No other age groups showed significant differences in loneliness levels. Results suggest that loneliness levels generally decreased with age.

Further chi-square analysis revealed significant associations between loneliness and both marital status (x^2^ (1) = 47.92, p < .001) and age (x^2^ (5) = 13.29, p = .021), with single individuals and younger adults reporting the highest rates of loneliness (48.4% and 43.8% respectively). Participants reported generally high life satisfaction levels, with a mean of 7.74 on a scale of 0-10 (SD=1.448), with only 2% reporting low life satisfaction compared to 10.6% in the general population[52]. Further analysis revealed a statistically significant difference in life satisfaction across age groups (f = 6.46, p < .001). Post hoc comparisons (Tukey HSD) indicated that participants aged 65 and over (M = 8.37, SD = 1.17) reported significantly higher life satisfaction than those aged 18–24 (M = 7.28, SD = 1.67, p = .006). Overall, the results suggest that life satisfaction tends to increase with age.

#### Promotion of Ireland Lights Up

The majority of participants learned about ILU from GAA club newsletters or GAA focused social media (n=656, 81.5%). A small number of participants learned about the initiative through national TV (n=46, 5.7%) or through family and friends (n= 38, 4.7%). The remaining participants (n= 39, 4.95%) heard about ILU through other means such as church newsletters, community pages in local newspapers, with a small number learning about it through the health insurance provider Irish Life (n=26, 3.3%). In general, participants were satisfied with the organisation of ILU, with a high mean score of 8.48 (±1.94), on a scale of 0-10.

### Qualitative Results

#### Drivers of Participation in Ireland Lights Up

The following section presents three major themes derived from interview and open-ended survey data, representing the key drivers of participation in ILU. Participants were asked who they participate with and why they participate, with the aim of identifying the factors that motivate engagement and to inform how demographic gaps might be addressed in future iterations of the initiative. Themes are supported with illustrative quotes, identified by participant number.

#### A Safe Place Where None Existed

Safety was identified as one of the strongest drivers of participation across the dataset, particularly salient among those living in rural communities. Participants consistently described the physical environment of rural Ireland in winter as a significant barrier to PA, characterising unlit, narrow roads and speeding traffic as deterrents to walking outdoors. For many participants, the provision of an enclosed, well-lit walking track at their local club was described, not as a convenience but as a fundamental enabler of participation, or *‘lifeline’* during the winter. One participant described ILU as:

> “a great initiative as it gives people a safe place to walk at night-time. Where I live it’s not safe as the roads are dark and lonely” (P56).

Other participants highlighted that in many cases, this was the only viable option for being physically active during winter evenings, with one participant explaining:

> “I can’t get out for a walk in the evenings when living in a rural area in the winter as it’s dark and the closest town is a 20mins drive away” (P85).

Participants consistently framed the absence of safe walking spaces not as merely an inconvenience but as a situation that made unsupported outdoor activity genuinely inaccessible. Another participant explained that the initiative *“gets people together for some light-hearted exercise and off the roads where you’re at risk of accidents in rural areas … narrow roads, poor if no lighting, no pedestrian paths” (P127),* positioning ILU not as one option among many, but as filling a structural gap in rural PA infrastructure. Notably, rural participants were more likely to cite safety and environmental accessibility as an important driver, suggesting this concern is geographically patterned.

Beyond the physical environment, participants described a further dimension of safety rooted in the collective experience of walking together in groups. For some participants, particularly women who would not walk alone after dark, the presence of others was described as the deciding factor in their participation.

> “I wouldn’t go walking in the village by myself – but in a group, you get the chat, the safety and the motivation.” (P368).

For some, this was the reason they felt comfortable taking part and for others, it was the only way they could be physically active in a safe manner, describing it as*“…only safe place in the village” (P542)*. Participants explained that knowing others would be present gave them the incentive to turn up. In this way, ILU was seen as both a catalyst for PA and enabled initial participation during the winter months. This social dimension of safety was distinct from the environmental one as it was not only about the track being lit but about not being alone on it.

Participants stressed the importance of ILU in offering a way to stay active when long, dark evenings might otherwise lead to inactivity. As one participant reflected, *“In rural areas in winter the nights are so long it can lead to a very sedentary lifestyle. But knowing there’s a safe place to go in the evenings to exercise is a great help” (P104).* This sense of reassurance extended beyond individual safety to the collective, with participants valuing the inclusive nature of ILU. Participants consistently emphasised the low-pressure, non-competitive nature of ILU as a further accessibility feature. The absence of long-term commitment or performance expectations was described as welcoming to people of varying ages and abilities. For some participants, the accessible environment was transformative: *“I walk with a stick, and I can walk safely around it … it’s fantastic!” (P70)*. ILU was described as the *“spark”* that encouraged people to get out and be active during the winter. In this sense, accessibility was understood by participants not only in physical or geographic terms, but as a characteristic of ILU’s culture and design by lowering the threshold for participation and making sustained engagement feel achievable.

#### Walking as a Mechanism for Social Connection

Participants consistently described ILU as more than just a walking initiative, recognising it as a driver of social interaction and community connection during the winter months. Across the data, participants characterised the initiative as creating a dedicated space for social engagement at a time of year associated with reduced PA, isolation and limited opportunities for community gathering. While PA was acknowledged as important, participants frequently positioned the social dimension as equally, or in some cases, more important to participation. Participants described ILU as a source of motivation, emphasising its role in supporting wellbeing and framed it as a tool not just for PA but also for supporting social and mental health.

> “… a great initiative to bring the community together and meet people you wouldn’t ordinarily meet, plus it is a great initiative to get us outdoors and exercising while meeting people too.” (P16).

> “the walks have fostered … a real feel of togetherness in what can be a very tough few months for people.” (P86).

The reference to winter as “a tough few months” was a recurring motif across participant accounts, suggesting that the timing of ILU was understood as intentional and meaningful. Participants did not simply describe enjoying the social aspects of the walks. Many articulated it as addressing a specific seasonal need, positioning ILU as a structured response to the social withdrawal and isolation that characterises this season for many people. A minority of participants who lived alone highlighted the importance of post-initiative social moments which allowed opportunity for face-to face interactions. Notably, participants in this group did not describe the walks primarily in terms of physical health benefits, but rather as reliable opportunities for social interaction.

> “…a great social initiative and the refreshments after in the club house were equally as important as the physical activity.” (P212).

This finding was not limited to those who lived alone. Across a broader range of participants, post-walk social moments were described as an integral component of the initiative rather than an optional extra, suggesting that the social infrastructure surrounding ILU was as valued as the walk itself. Participants also framed ILU as a catalyst to reconnect with each other and get back to PA after the holiday season. Participants observed that it created a reason to re-engage with their local club while the playing season was paused.

> “a brilliant way to meet up post-Christmas to walk it off… lovely to be back at the pitch after the winter break” (P136).

Other participants noted that in the post-pandemic world, the ILU initiative offered a space to get active, be together and unite communities. The reference to the post-pandemic context appeared across a subset of accounts, with these participants situating ILU within a longer experience of social disruption and loss of community. For this group, the initiative represented not simply a new initiative but a means of rebuilding social habits and connections that had been interrupted.

> “Covid-19 destroyed everything – people are lonely – this helps so many people to get out again and meet up. It’s fantastic!” (P80)

This sense of social connection was not limited to existing networks; the walks also fostered new relationships that transcended age and backgrounds and that might not otherwise have formed. For those new to an area, ILU provided a valuable entry point into local life, described as a way for newcomers to get to know others in the community and highlighting the wider social value for clubs and members.

> “ILU gets people from all ages walking and chatting together and creates a real sense of community and belonging.” (P46).

Some intercultural interactions showed the potential for inclusivity with a small number of participants remarking that participating in ILU provided a way to break down barriers, as people from different cultures were able to meet and interact in a safe space. However, it is important to note that these interactions were uncommon and the exception rather than the rule as the majority of participants were from a local Irish background. While some accounts pointed to intercultural interaction, these were relatively rare and should not be interpreted as indicative of broader inclusivity.

> “a great way of meeting people … from different cultures … last week I walked with a Brazilian couple, and we had great conversations regarding our different cultures.” (P24).

Th multigenerational character of ILU was a distinct and valued feature across accounts. Participants described the initiative as one of the relatively few structured opportunities for different age groups to engage together in a shared activity. The initiative offered a way to get children outside and spend time together in the safe, well-lit club setting. Likewise, the initiative offered an opportunity and was accessible for families to bring along children of all ages to participate and in several cases, participants described multi-generational family groups attending together, positioning ILU as an event for all age cohorts. This suggests ILU appeals to family-based and intergenerational participation, potentially shaping the demographic profile observed.

> “a great initiative to get families out walking together. My children love to go on the walk and meet their friends” (P23).

> “it’s a social activity for all ages. I bring my elderly parents with me, and we love the exercise and connections the event brings.” (P116).

#### From Sporting Venue to Community Asset

The third theme highlighted another driver of participation: the role of both walking track infrastructure and social infrastructure within clubs. While ILU was often described as the initial draw, participants emphasised that the walking tracks themselves and the social connections they fostered were what encouraged people to keep coming back. The tracks were characterised as busy community spaces throughout the day, with both participants and club leaders framing the constant activity and usage they generated as the primary marker of success.

> “The track is fantastic for the village and ILU has kicked it off in a great way – it’s like the hook to promote it and tells people, ‘Come in – this is for you’!” (P289).

> “… if you stand here from 7am to 9pm, you’ll see the track is used all day long – that’s the success for the club.” (P55).

Notably, participants did not describe the track’s value in terms of PA alone. Instead, sustained usage was interpreted as evidence of community ownership and belonging, with the track functioning as a visible sign that the club had become meaningful to a wider group of people. This was particularly evident among participants who also volunteered in a leadership capacity as they described footfall and activity as indicators of the initiative’s wider community impact.

The infrastructure also served to bridge gaps between past and present members, sustaining ties to the club through accessible physical space. Describing the track as a low-threshold re-entry point into club life, participants pointed to the wider benefits for involved clubs, from raising their profile to attracting new members and making full use of facilities and walking tracks during quieter periods in the sporting calendar.

> “… for people who spent many years on a committee, then finish, they can lose that connection, so this is a link to the club and the community.” (P431).

> “… this time of year, is ideal for ILU because it’s a dead time for the club … but ILU brings people up as a focus for the community.” (P237)

These accounts suggest that participants understood the track not simply as a facility, but as a means of maintaining social identity and connection to the club across different life stages. The quotes further illustrate how participants positioned ILU as serving an organisational function in filling a structural gap in the club calendar and providing PA opportunities during quieter periods of sporting activity.

Importantly, ILU, as a way to promote club walking tracks, was seen as a small shift towards greater inclusivity, helping to open GAA clubs to people who might previously have felt disconnected from the organisation. This was not framed as a programmatic outcome alone, but as a signifier of a wider cultural shift within clubs, with several participants drawing an explicit contrast between the club’s historical identity and its present orientation.

> “GAA clubs traditionally came with high walls… something’s changed. This initiative is a great example of that new approach to whole community involvement.” (P197)

This contrast between past exclusivity and present openness appeared across the dataset, suggesting that for many participants, engaging with ILU carried a symbolic dimension beyond PA, where the tracks and ILU as a social outlet were valued not only for supporting walking, but as a means of redefining the role of the local club, positioning it as a viable space for health, wellbeing and whole-community involvement. Collectively, these findings suggest that participation in ILU is shaped by a combination of environmental, social, and infrastructural factors, which may also influence the types of populations reached by the initiative.

## DISCUSSION

The aim of this paper is to understand participation drivers and patterns by examining the demographic characteristics that influence participation, identifying those not reached, and exploring how participation is shaped by the cultural context. ILU is a low-cost, nationwide, sports club-based community walking initiative that is unique in its scale and reach. Nearly half of all GAA clubhouses hosted ILU walking sessions, with 784 clubs registered to participate. Determining the overall reach of ILU was challenging due to the lack of centralised collation of weekly participant registration data and the overlap of multiple clubs using shared facilities. Retention rates were positive with a consistent and engaged group returning each year. Previous literature has shown that walking groups attract large numbers of participants, as evidenced by community walking initiatives such as Australia’s Heart Foundation Walks [20] and 10,000 Steps Ghent [53]. As shown by the reported results, ILU demonstrates substantial national reach. However, it is important to recognise that reach is not merely about the number of participants but also about who participates and who remains unreached. Understanding these patterns is central to evaluating the initiative’s impact and informing strategies to engage not yet reached groups.

In terms of socio-demographic characteristics, ILU participants were predominantly female, White and GAA-affiliated, living in mainly rural areas within 5 kms of their club. The majority of participants held third level qualifications and were aged between 35 and 54 years. Most participants learned about ILU through GAA club newsletters or social media. The results highlighted that participants had high levels of health and wellbeing with strong life satisfaction, sense of belonging and adherence to PA guidelines. This demographic profile of participants reveals both strengths and weaknesses in the current reach of ILU. The initiative predominantly engages a mid-life, well-educated, female population. This finding is supported by broader evidence that women are more likely than men to engage in organised walking and community health initiatives [14]. In the Heart Foundation Walks, participants tended towards a similar demographic profile, with an average age of 64 and 80% being female [20]. Irish national data shows a 7% gender gap in recreational walking favouring women [54]. These findings suggest that the social, non-competitive format of walking appeals differently to men and women or that men connected to GAA culture associate the organisation primarily with competitive sport rather than recreational PA [14, 55]. Interestingly, while researchers and public health authorities have increasingly recognised the potential of sports clubs as vehicles for health promotion, the evidence base remains limited in important respects. A recent systematic review found that health promotion interventions delivered within sports club settings targeted predominantly male participants in team sports [15]. Therefore, walking groups in this context may function as feminised health spaces built around social connection and the absence of competitiveness, features that align with the gendered roles often observed within GAA clubs. Men may be reluctant to join walking groups due to fear of being in a minority, compounded by HP messaging around walking that has historically targeted women more directly [14]. This suggests that GAA affiliation alone does not ensure reach; rather, the nature of the activity, the environment and the social interaction itself shapes appeal.

Nonetheless, an intervention can’t always be tweaked to appeal to everyone. The fact ILU appeals to women is very positive as although more females participate in walking, females of all ages are less active than males [54]. Women, in particular, face significant barriers to PA participation, especially in rural and underserved communities, where challenges related to access, safety, social norms and competing life demands can limit engagement [13]. Against this backdrop, the present study is notable in that it successfully engaged primarily women, a population that is consistently under-represented in both sports settings and health research more broadly [15]. Women remain under-represented in many sports settings, including as research participants. This was evident in a recent scoping review of research on women in sport which found that in Ireland between 2014 and 2020, only 34% of participants in sport and exercise science studies were female [56]. That this initiative reached and retained women, particularly in a rural community setting, is therefore a meaningful finding in itself and one that warrants careful consideration of the factors that enabled their engagement. Participation was closely linked to the safety ILU provided, both the secure, well-lit environment and the social reassurance of walking groups, addressing a key barrier to PA in this context

Participation rates were modest among younger adults (3% aged 18-24), despite findings indicating that younger adults were more likely to report being lonely. While this may be due to the initiative’s format and timeslot, club-based community initiatives may be perceived as culturally distant for a generation who are more likely to be engaged in competitive sport and whose social connections increasingly form through technology-based mechanisms [57]. Notably, married or cohabiting participants, and GAA members, reported lower loneliness levels, reinforcing the protective role of close relationships and community connections [58–61]. As older adult participants reported reduced loneliness levels, there may be opportunities to implement targeted approaches highlighting the possible benefits from participating in ILU to younger cohorts. Participants generally reported high wellbeing, with strong life satisfaction, sense of belonging and adherence to PA guidelines. While causal inferences cannot be drawn from the quantitative data, the psychosocial drivers of participation were particularly evident in the qualitative data, with participants reporting a strong sense of belonging to their community and club. Many described the initiative as a source of routine, social connection and accountability. These enablers are consistent with behaviour change techniques which support sustainable PA levels, such as accountability, social support and environmental cues [62, 63]. The findings also support previous literature which show the role of structured, time-bound, place-bound initiatives in enhancing social capital, fostering trust through sustained and localised connections [64]. Similarly, the multi-generational nature of ILU was identified as a strength. Evidence from intergenerational exercise interventions point to positive impacts on both younger and older participants, including psychosocial wellbeing and motor skills [65]. Parents valued the safe space for children to play while they walked, while others highlighted the benefits of shared participation across age groups.

High rural uptake is a defining feature of the ILU initiative. This reflects the geographical footprints of GAA clubs, many of which serve as the fulcrum for rural communities where volunteers fulfil multiple roles to bring the community together [66]. From a public health perspective, this reach is significant as rural populations are traditionally ‘hard to reach’, often facing isolation, limited infrastructure and fewer opportunities for safe PA [67, 68]. By offering accessible, floodlit walking tracks, ILU addresses a practical barrier to walking in rural Ireland: the absence of safe, lit footpaths. The ‘localness’ of the clubs as an enabler to communities means that ILU is valuable during winter, when safety concerns and darkness restrict outdoor activity but also as a potential ‘hook’ to increase PA levels year-round by using the club as a setting for HP. The initiative demonstrates the importance of environmental infrastructure in enabling population PA, particularly in geographically isolated areas [69].

The sample was notably homogeneous in terms of education and ethnicity. Over 80% of participants held third-level qualifications, compared to 56% of the Irish adult population [70]. This reflects well-established links between education and positive health behaviours [71, 72] but has implications for tackling health inequities. The absence of ethnic diversity may reflect both the demographic profile of rural Irish communities and past historic insularity of GAA clubs [73, 74]. For ethnic minority or immigrant communities, GAA clubs may represent unfamiliar spaces with unclear expectations. Language barriers, cultural unfamiliarity with the GAA’s role in Irish life and inbuilt social networks may contribute to a lack of awareness and thus, non-participation.

In considering these findings, it is important to reflect on the social meaning of walking. Walking itself is accessible, low-barrier and free, yet organised sports club-based walking carries specific cultural meanings and social expectations that shape participation in unequal ways [75]. As discussed, walking as a form of PA may hold greater appeal to certain groups over others. Consideration needs to be given to positioning walking as a legitimate and socially valued form of PA by emphasising the benefits of social connection and wellbeing [76]. Yet, even if certain groups are proportionally underrepresented, ILU’s large overall scale means that it still engages a considerable absolute number from subgroups. There is room to celebrate its success while working strategically to broaden reach. However, careful consideration must be given to whether expanding reach is possible within the culture of the GAA whose primary focus is to promote Gaelic games or instead positioned as a catalyst for involving more community members in walking to form new long-lasting habits. Expanding reach will require more deliberate outreach and diverse communication channels with efforts made to ensure that these strategies are genuine and not tokenistic.

Current communication and promotion strategies may unwittingly reinforce existing participant characteristics. As most participants reported hearing about ILU through GAA promotional sources, it indicates that although it is a community initiative at scale, it is likely not extending the reach significantly beyond the GAA community. The GAA’s unique identity, whilst often an enabler to PA, may also constrain reach by primarily appealing to those who identify with the organisation. There is a risk of ‘preaching to the converted’ as most participants already associated with a sports club fulfil the role of highly motivated individuals, which the literature indicates are the groups most likely to benefit from such PA initiatives [77, 78]. Research indicates limited effectiveness of PA campaigns that rely solely on a single channel or approach tied to a specific organisation [77, 79, 80]. Similarly, by assuming existing familiarity with the GAA, promotional messaging that uses cultural references and channels specific to that community are unlikely to resonate with those outside it. Communication channels for ILU may not have penetrated beyond club members. This runs the risk of exacerbating existing disparities and underscores the importance of accounting for social inequities when designing and implementing mass PA campaigns [81, 82].

To effectively extend the initiative’s reach beyond GAA members, strategies must focus on diversifying message content, tailoring delivery and ensuring external engagement and acceptability [83, 84]. Message content should be designed to be relevant to specific groups rather than relying only on traditional demographics [82]. It is important to understand why certain interventions appeal to some groups and not others. Strategies to deliver messages through other credible organisations, peers and health care professionals should be considered [82, 85]. Finally, in order to expand reach, it is essential to involve the target audience in message development, by bringing together a variety of stakeholders that bring about meaningful changes and avoid tokenistic claims of inclusivity [86].

### Strengths and limitations

The study utilised a robust mixed-methods implementation science design, integrating quantitative surveys and qualitative snapshot interviews. Conducting snapshot interviews in situ allowed researchers to capture immediate perceptions and contextual insights that retrospective interviews often miss. However, their brief, in-situ nature may limit the depth achieved through longer, retrospective semi-structured interviews. Researcher presence during walks may have influenced how participants described their experiences. Additionally, the research team comprises academics with backgrounds in health promotion. While the team employed critical reflection and triangulation to mitigate this, their interpretive lens may still have influenced the thematic analysis of qualitative data.

The evaluation was guided by the RE-AIM and PRISM frameworks, ensuring systematic assessment of reach and adoption. However, non-random convenience sampling in recruiting clubs and participants may have introduced self-selection bias, with more health-conscious individuals choosing to take part. As a result, findings may not reflect a representative cross-section of participants across all GAA clubs in Ireland. Similarly, quantitative results rely on self-reported measures of PA levels, health status, and loneliness. Such data are susceptible to social desirability bias and recall errors, potentially leading to an overestimation of healthy behaviours.

## CONCLUSION

Sports clubs have been identified as suitable settings for community PA initiatives, yet this is one of the first studies to capture the demographic characteristics of a nationwide sports club-based PA intervention operating at scale. The findings demonstrate that ILU is already well placed to reach a large cohort of the population to improve PA levels and wellbeing across Ireland. Notably, ILU addresses two important populations often underserved by PA initiatives: rural populations experiencing social isolation and limited safe, well-lit infrastructure, and women who have lower PA levels than men. The provision of accessible, floodlit walking facilities during the winter months may help reduce social and infrastructural barriers to participation in PA. The initiative’s success demonstrates its effectiveness as an easily implemented, scalable model with significant public health reach.

While there remains scope to broaden participation among other demographic groups, ILU’s current reach to priority populations is substantial. Future research should give attention to implementation factors and evaluate longer-term impacts on participant health outcomes to support ILU’s place within national policy, guiding roll-out to other National Governing Bodies of Sport. From an international perspective, ILU provides a rare example of national-scale, sports club-based PA intervention. Lessons from its successes may be transferable to other sports settings globally, particularly those with strong community infrastructure and popular indigenous sports.

## Abbreviations

PA: Physical Activity
ILU: Ireland Lights Up
GAA: Gaelic Games Association
ISRCTN: International Standard Register for Controlled Trials Number
RE-AIM: Reach, Effectiveness, Adoption, Implementation, Maintenance
PRISM: Practical Robust Implementation and Sustainability Model

## Acknowledgements

The authors wish to thank the GAA and GIW for supporting the project and the individual GAA clubs, club leaders and participants for participating in and supporting Ireland Lights Up.

## Statements and Declarations

### Authors’ contributions

AM, BL and NB were responsible for the study design. NB was responsible for writing the original draft and substantive review and editing. AM, BL, NM and NR provided supervision and substantive review and editing. All authors read and improved the final manuscript.

### Availability of data and materials

The profile of this research project will be housed within the Centre for Health Behaviour Research in SETU and data will be stored for 10 years post publication of outcomes. Anonymised datasets from this study will be made available (as allowable according to institutional IRB standards) by emailing the corresponding author.

### Ethics Statement

The study received ethical approval from South East Technological University Health Sciences Research Committee (SETU/HSREC/23/24/010). This study has also been registered with the ‘International Standard Randomised Controlled Trial Number’ registry (ISRCTN 14693503).

